# Comprehensive knowledge and positive attitudes regarding HIV/AIDS among reproductive-aged women in Bangladesh and their associated factors: a cross-sectional study using the Multiple Indicator Cluster Survey 2019

**DOI:** 10.1101/2024.03.06.24303887

**Authors:** Md Arif Billah, Raba’Aton Adawiah Mohd Yusof, Md. Nuruzzaman Khan, Ruhani Mat Min

## Abstract

**Background:** HIV/AIDS remains a global health concern and understanding the knowledge and attitudes of at-risk populations is crucial for effective prevention and management. This study examines the knowledge and attitudes related to HIV/AIDS among reproductive-aged women in Bangladesh and explores associated factors.

**Methods:** The study analyzed data from a total of 39,066 reproductive women, obtained from the nationally representative 2019 Multiple Indicator Cluster Survey. The outcome variables assessed were the level of HIV/AIDS-related knowledge (high *vs* low) and attitudes regarding HIV/AIDS (good *vs* poor). These variables were derived by calculating the median values of scores obtained from nine and six questions in the survey that assessed knowledge and attitudes toward HIV/AIDS, respectively. The explanatory variables included sociodemographic factors and variables related to media exposure. A multivariate logistic regression model was employed to investigate the associations between the outcome and explanatory variables.

**Results:** Over half of the total respondents (51.6%) reported a lower level of knowledge, and a significant majority (54.2%) exhibited poor attitudes regarding HIV/AIDS, with notable variations observed across districts. Women in comparatively higher age groups, those with higher levels of education, hailing from more affluent families, residing in urban areas, and having exposure to newspapers, the internet, computers, and mobile phones displayed higher levels of knowledge regarding HIV/AIDS. Conversely, women with higher education levels, residing in rural areas, and having exposure to mobile phones and the internet exhibited good attitudes regarding HIV/AIDS. Unmarried respondents reported lower likelihoods of good attitude towards HIV/AIDS.

**Conclusion:** The findings underscores the urgent need for policies and programs designed to improve HIV/AIDS-related knowledge and attitudes, with a focus on district-level requirements. Effective strategies may include mass media campaigns and the incorporation of HIV/AIDS- related topics into the national curriculum.

## Background

Since 1981, HIV/AIDS has emerged as a prominent and enduring global epidemic. The global prevalence of HIV stands at approximately 39.0 million individuals at the end of 2022, with an estimated range of 33.1 to 45.7 million. Notably, among this affected population, approximately 38.5 million are adults aged 15 and above, signifying a substantial public health burden [1]. In the year 2022 alone, more than 1.3 million individuals experienced new HIV infections, with females accounting for 49% of these cases [1]. It is worth mentioning that the regions of Asia and the Pacific occupy the second position in terms of HIV prevalence, with roughly 6 million individuals living with HIV, trailing behind sub-Saharan Africa [2, 3].

In the context of Bangladesh, an Asian nation with a Muslim-majority population and ranking as the 8^th^ most populous country globally, the prevalence of HIV/AIDS remains relatively low, with a total of 15,000 diagnosed cases thus far [4]. A significant portion of these individuals are presently integrated into the government-provided healthcare service network, with 4,565 individuals receiving antiretroviral therapy (ART) treatment [4]. Nevertheless, despite this lower prevalence, Bangladesh has experienced two notable increases in HIV incidence, one in 2015 and another in 2021, during which a total of 729 new cases were reported, with women accounting for 22% of these cases [5]. The newly reported cases predominantly consist of young individuals, with 96% falling within the age range of 15 and over years, including 16% from 15–24 years, 67% from 25–49 years, and 13% of them from 50+ years; while a majority of them are married (60.4%) [5]. Furthermore, there has been a proportional increase in HIV/AIDS-related deaths, with the highest number of such deaths (n=205) recorded in the year 2021 [5].

The growing trend in HIV prevalence and its geographic dispersion, the ongoing high-risk behaviours of drug users and sex workers, and the significant mixing of the most at-risk groups with the bridging populations serve as reminders that HIV may not stay isolated in any one neighbourhood or subpopulation [5–8]. Alongside, about 46% of the HIV incidence occurred due to cross-border migrations [5], including forced migrants from Myanmar, i.e., the Rohingya population. These populations also elevated the prevalence of HIV/AIDS and other STIs in Bangladesh [9–11]; by integrating drug use and sex worker business with Bangladeshi locales [9, 12, 13]. Bangladesh provides a huge labour market supply to developed countries, where the stigma and lack of HIV screening upon arrival and departure of those migrants caused an increased level of HIV transmission [14].

In all cases, women are considered the most vulnerable group due to traditional customs, power dynamics, and sexual and reproductive health practices [15–17]. All these hinder women from negotiating protective measures, such as condom use, which further exacerbates this challenge [15, 18–22]. They are at greater risk of HIV/AIDS transmission due to associated socioeconomic exposures compared to men [23]. Moreover, it become worse because of persistent gender inequality and human rights violations [24]. In the last behavioural surveillance-2016, 5% of the female people who inject drugs (PWID) were recorded to have HIV infection; indications of HIV transmission are beginning to spread to female PWID [8]. This underscores the urgent need for targeted interventions and awareness campaigns aimed at preventing HIV/AIDS transmission as well as improving access to care and treatment. However, in order to effectively address these challenges, it is crucial to have a comprehensive understanding of the knowledge and attitudes regarding HIV/AIDS among the reproductive-aged women in Bangladesh.

There is a significant gap in the studies identifying the levels and factors, specifically the level of attitudes and geographical variations of knowledge and attitudes among the women in Bangladesh. These studies primarily focused on HIV/AIDS knowledge and its associated sociodemographic factors [23, 25–32]. According to these studies, women with higher levels of education, residing in urban areas, being exposed to mass media, working outside the home, and higher wealth quintile were played important roles in raising awareness about HIV/AIDS. Moreover, comprehensive knowledge items were often treated as separate components from sociodemographic factors [31].

However, these studies ignores the other online media which also plays a vital role in health literacy and disease prevention. In comparison to HIV/AIDS knowledge, there are few studies examining HIV/AIDS attitudes, which have primarily explored discrimination and stigmatization among the general population [17, 33–35], as well as among healthcare professionals [33, 36, 37]. Few studies had explored the attitudes of HIV patients, of which internal stigma and shame were common to them [17, 34]. While these stigmas and discriminatory attitudes were considered critical factors for the national approach to HIV prevention [38]. Importantly, these studies mostly overlook regional variations in HIV/AIDS prevalence, which are found to be higher in both Bangladesh and other LMICs.

Therefore, a lack of evidence on both HIV/AIDS-related knowledge and attitudes can create challenges in terms generalizability of the findings for vulnerable women, which is important for policymaking and program development. To address these gaps, this study was conducted to explore knowledge and attitudes regarding HIV/AIDS among reproductive-aged women in Bangladesh, while also examining regional-level variations and associated factors.

## Methods

### Sampling and data

The data for this study were obtained from the 2019 Bangladesh Multiple Indicator Cluster Survey (MICS), conducted by the Bangladesh Bureau of Statistics (BBS) in collaboration with UNICEF from January 19 to June 1, 2019. The survey employed a two-stage stratified random sampling procedure to select nationally representative households from which data were collected. In the first stage of sampling, a total of 1,300 Primary Sampling Units (PSUs) (areas with an approximate population of 200 to 300 households) were selected. In the second stage, 20 households were chosen from each PSU using systematic random sampling techniques, resulting in a total sample of 64,400 households. Among the selected households, 61,602 were occupied, and of these, 61,242 were successfully interviewed, resulting in a household response rate of 99.4%. Among the selected households, a total of 68,711 eligible women met the inclusion criteria: these women either resided in the selected households as usual residents or had stayed there during the most recent night. Of these eligible women, 64,378 were successfully interviewed, yielding a response rate of 93.7%. Data were collected using a pre-structured and pre-tested questionnaire implemented in all MICS surveys conducted throughout LMICs. This questionnaire captured insights on various aspects, including fertility, early childbearing, family planning, unmet need, antenatal care, neonatal tetanus, delivery care, birthweight, postnatal care, maternal morbidity, as well as knowledge and attitudes related to HIV/AIDS. For additional details about the sampling procedure, please refer to the final report of the 2019 MICS [39].

### Analytical sample

The sample for this study was selected based on two criteria: (i) women aged 15-49 years and (ii) respondents who provided their responses to HIV/AIDS knowledge and attitude-related items during the survey. Initially, the survey inquired, “Have you ever heard about HIV/AIDS?” Respondents who answered affirmatively were then presented with knowledge and attitude-related items (see supplementary table 1). Initially, 39,009 women responded positively to the first question and were considered eligible. However, 487 of them did not respond to the knowledge and attitude-related items and were subsequently excluded from the analysis. Finally, 38,522 women were included in the analysis (see the supplementary Fig 1 file).

### Outcome variables

Two outcome variables were considered: (i) Level of comprehensive knowledge regarding HIV/AIDS (high, low) and (ii) Level of positive attitudes towards HIV/AIDS (good, poor). The survey included a total of nine questions for measuring knowledge and six questions for attitudes. The themes of these questions are presented in Table 2 and 3 for knowledge and attitude items, respectively. The exact questions asked can be found in Supplementary Table 1 and 2. Respondents were given three response options for each question: “yes,” “no,” and “don’t know” for knowledge items and “yes,” “no,” and “don’t know/not sure/depends” for attitude items. We recategorized these responses as 1 for a correct answer and 0 for a wrong answer or if the answer was “don’t know.” This generated a continuous score ranging from 0 to 9 for knowledge items and 0 to 6 for attitude items, where a higher score indicated greater knowledge and more positive attitudes. Subsequently, we calculated the median scores for knowledge (median=6) and attitude items (median=3) and reclassified respondents as having a high level of comprehensive knowledge and good level of positive attitude regarding HIV/AIDS if their individual score was equal to or greater than the median score.

### Explanatory variables

The selection of explanatory variables in this study occurred in three stages. Firstly, an extensive literature search was conducted across various databases to compile a list of variables considered in prior studies within LMICs [23, 26, 29, 30, 40–54]. Secondly, the survey’s dataset was examined to verify the availability of these variables. Subsequently, the statistical significance of the available variables concerning the outcome variables was assessed in the third stage, and only those that displayed statistical significance were included in the final analysis. These selected variables encompassed respondents’ age (15-19, 20-24, 25-29, 30-34, 35-39, 40-44, 45-49), respondents’ education (no education, primary, lower secondary, higher secondary, and higher) and respondents’ marital status (never married, currently married, widowed, divorced, separated). The variable of household wealth quintile, derived by MICS through principal component analysis of household assets, such as ownership of radios or televisions, was also incorporated. Additionally, factors related to media exposure were considered, including reading newspapers or magazines (yes, no), listening to the radio (yes, no), watching television (yes, no), using a computer (yes, no), using the internet (yes, no), and using a mobile phone (yes, no). Geographical factors were accounted for by considering place of residence (urban and rural) and regions (Barishal, Chattogram, Dhaka, Khulna, Mymensingh, Rajshahi, Rangpur, Sylhet).

### Statistical analyses

Descriptive statistics were employed to elucidate the characteristics of the respondents, with the percentage distribution of each item pertaining to comprehensive knowledge and positive attitudes being reported. However, we also examined the skewness of these dependent variables (see supplementary Figs 2 and 3). A geographical distribution was also presented to examine the geographical variability of these variables. Multivariate logistic regression analysis was applied to investigate the factors associated with a high level of knowledge regarding HIV/AIDS and good attitudes regarding HIV/AIDS. Two distinct models were employed: one for assessing a high level of knowledge regarding HIV/AIDS and another for evaluating good attitudes regarding HIV/AIDS. Prior to executing each model, we conducted a thorough check for multicollinearity. If evidence of multicollinearity was detected, signified by a variance inflation factor (VIF) exceeding 5%, the pertinent variable was removed, and the model was re-run. Furthermore, we took into account the complex survey design inherent to the MICS survey, which encompasses strata, primary sampling units, and women’s sample weights [55]. For all statistical analyses, we utilized STATA software (Stata Corp, TX), while Arc GIS version 10.4 was employed to explore geographical variations.

## Results

### Sample characteristics

Table 1 provides an overview of the background characteristics of the respondents. Notably, a substantial portion of women fell within the teenage bracket (15–19 years, 21.7%). Furthermore, a significant proportion had completed their higher secondary education (46.4%), and the majority of the sample were in a current marital status (76.0%). Regarding socioeconomic factors, a noteworthy share of women belonged to the richer wealth quintile (28.9%). Geographically, approximately 72.5% of the total respondents resided in rural areas, with one-fourth (26.3%) situated in the Dhaka region. In terms of media exposure, a substantial majority reported watching television (77.4%) and using mobile phones (77.1%). Moreover, 18.4% had access to the internet, 13.3% engaged in reading newspapers or magazines, 7.2% utilized computers, and 3.9% tuned in to the radio.

**Table 1.**
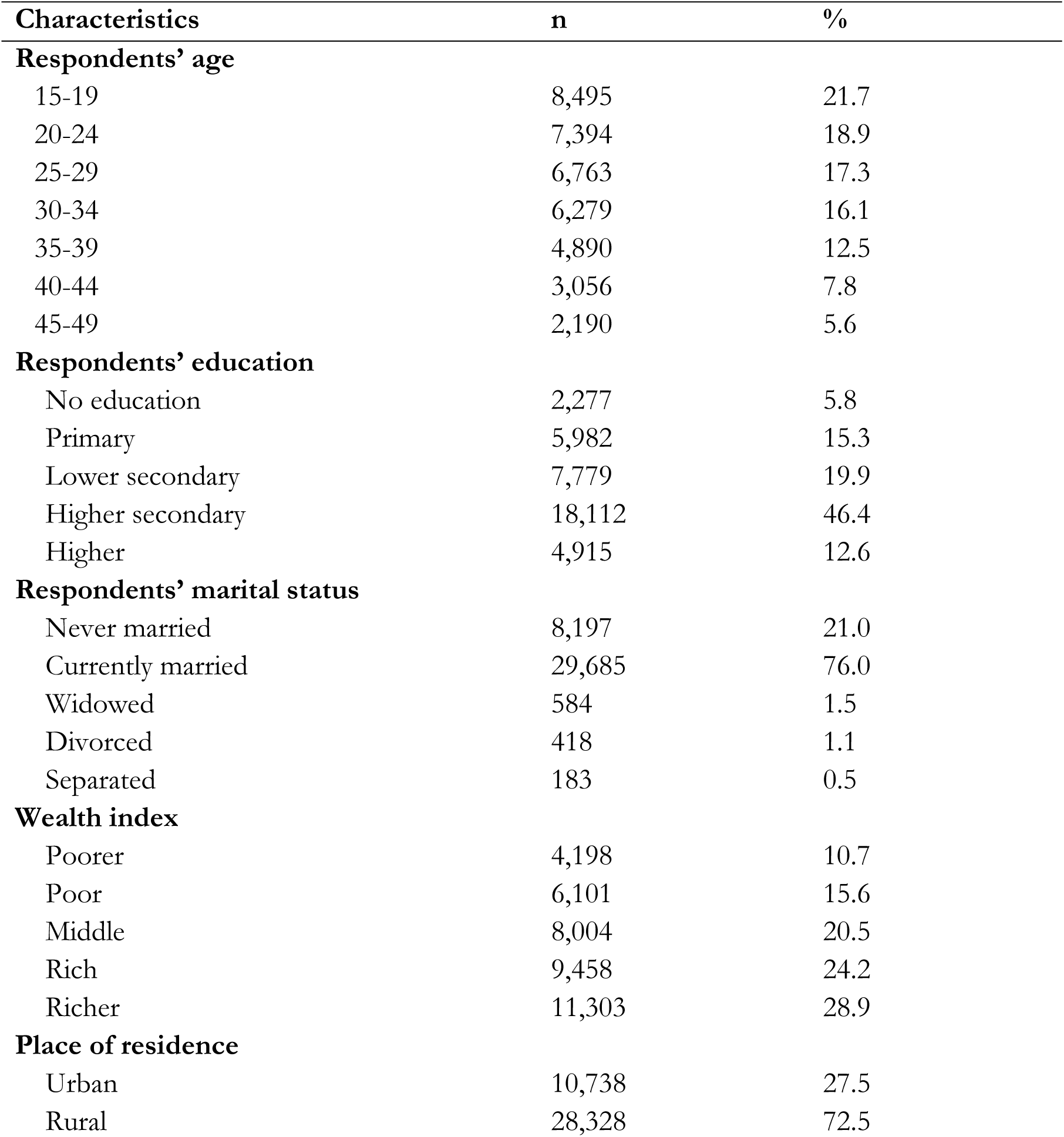
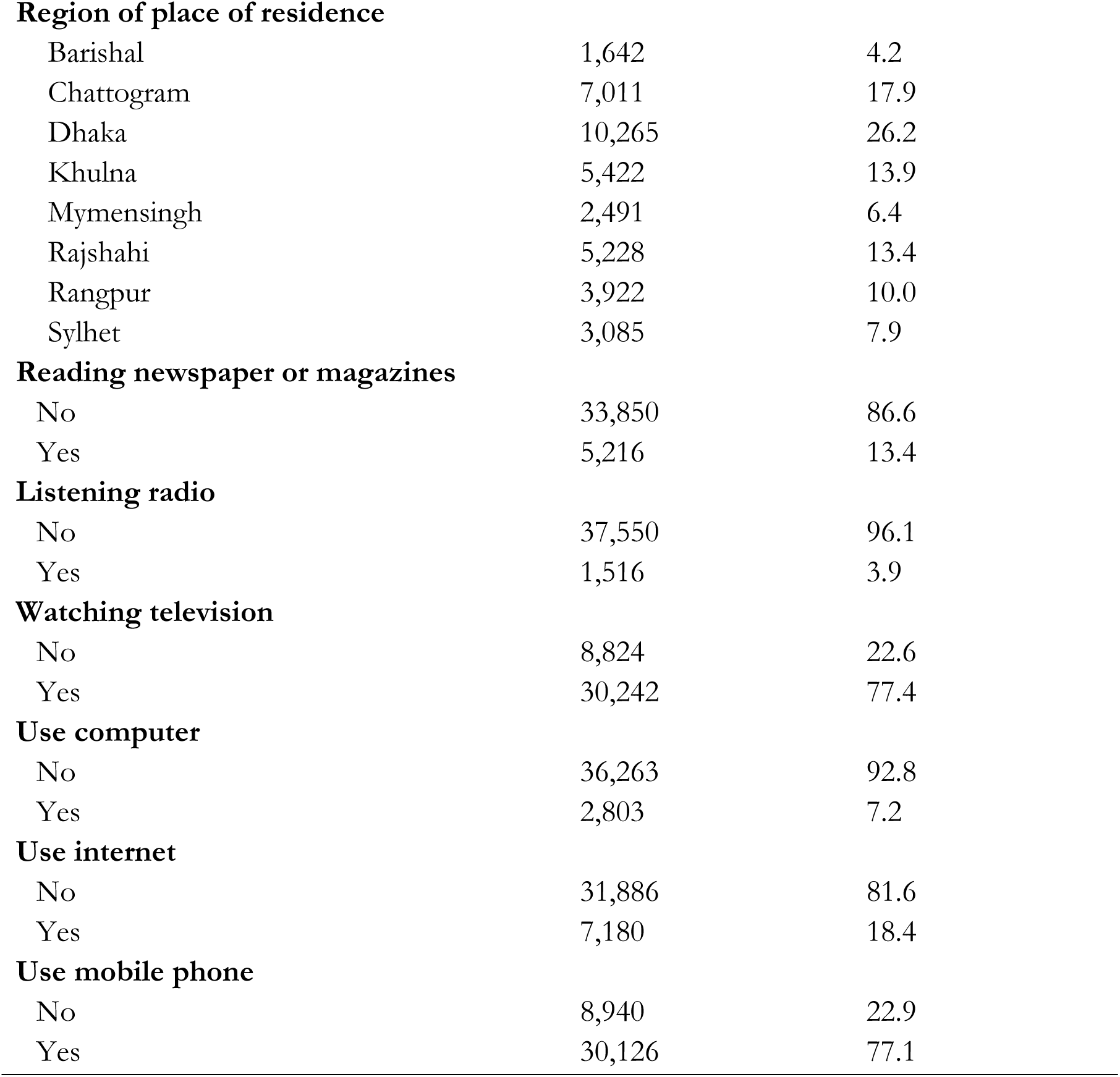
Sociodemographic characteristics the respondents, Bangladesh (N=39,066)

| Characteristics | n | % |
| --- | --- | --- |
| **Respondents' age** |  |  |
| 15-19 | 8,495 | 21.7 |
| 20-24 | 7,394 | 18.9 |
| 25-29 | 6,763 | 17.3 |
| 30-34 | 6,279 | 16.1 |
| 35-39 | 4,890 | 12.5 |
| 40-44 | 3,056 | 7.8 |
| 45-49 | 2,190 | 5.6 |
| **Respondents' education** |  |  |
| No education | 2,277 | 5.8 |
| Primary | 5,982 | 15.3 |
| Lower secondary | 7,779 | 19.9 |
| Higher secondary | 18,112 | 46.4 |
| Higher | 4,915 | 12.6 |
| **Respondents' marital status** |  |  |
| Never married | 8,197 | 21.0 |
| Currently married | 29,685 | 76.0 |
| Widowed | 584 | 1.5 |
| Divorced | 418 | 1.1 |
| Separated | 183 | 0.5 |
| **Wealth index** |  |  |
| Poorer | 4,198 | 10.7 |
| Poor | 6,101 | 15.6 |
| Middle | 8,004 | 20.5 |
| Rich | 9,458 | 24.2 |
| Richer | 11,303 | 28.9 |
| **Place of residence** |  |  |
| Urban | 10,738 | 27.5 |
| Rural | 28,328 | 72.5 |
| **Region of place of residence** |  |  |
| Barishal | 1,642 | 4.2 |
| Chattogram | 7,011 | 17.9 |
| Dhaka | 10,265 | 26.2 |
| Khulna | 5,422 | 13.9 |
| Mymensingh | 2,491 | 6.4 |
| Rajshahi | 5,228 | 13.4 |
| Rangpur | 3,922 | 10.0 |
| Sylhet | 3,085 | 7.9 |
| **Reading newspaper or magazines** |  |  |
| No | 33,850 | 86.6 |
| Yes | 5,216 | 13.4 |
| **Listening radio** |  |  |
| No | 37,550 | 96.1 |
| Yes | 1,516 | 3.9 |
| **Watching television** |  |  |
| No | 8,824 | 22.6 |
| Yes | 30,242 | 77.4 |
| **Use computer** |  |  |
| No | 36,263 | 92.8 |
| Yes | 2,803 | 7.2 |
| **Use internet** |  |  |
| No | 31,886 | 81.6 |
| Yes | 7,180 | 18.4 |
| **Use mobile phone** |  |  |
| No | 8,940 | 22.9 |
| Yes | 30,126 | 77.1 |

### Comprehensive knowledge and attitudes regarding HIV/AIDS

For comprehensive knowledge, it’s notable that nearly 48% of the total respondents reported a high level of knowledge regarding HIV/AIDS (Table 2). In terms of individual components of knowledge items, over two-thirds of the respondents acknowledged that HIV can be prevented by having an uninfected partner and consistently using condoms during sexual intercourse. Moreover, approximately 59.0% of the total respondents believed that a person who appears healthy can still carry the HIV virus. On the other hand, one-third of the respondents held the misconception that mosquito bites can transmit HIV from an infected person to a non-infected person. In the context of mother-to-child transmission, 73.0% of respondents agreed that HIV can be transmitted from mother to child during pregnancy, while 75.5% recognized the risk during breastfeeding. Furthermore, 57.3% of respondents acknowledged the potential transmission of HIV from mother to child during delivery.

**Table 2:** Percentage distribution of items measuring respondents’ HIV/AIDS knowledge and composite of comprehensive knowledge level (n=39,066)

| Knowledge Assessment Items for HIV/AIDS | No<br>% | Yes<br>% |
| --- | --- | --- |
| Can avoid HIV by having one uninfected partner | 32.1 | 67.9 |
| Can get HIV from mosquito bites | 68.8 | 31.2 |
| Can avoid HIV by using a condom correctly every time | 38.1 | 61.9 |
| Can get HIV by sharing food with person who has HIV | 62.7 | 37.3 |
| Can get HIV through supernatural means | 95.9 | 4.1 |
| Healthy-looking person may have HIV | 41.0 | 59.0 |
| HIV from mother to child during pregnancy | 27.0 | 73.0 |
| HIV from mother to child during delivery | 42.7 | 57.3 |
| HIV from mother to child through breastfeeding | 24.5 | 75.5 |
| Level of HIV/AIDS knowledge | Prevalence (95% CI) |  |
| Low | 51.6 (95% CI, 50.9–52.3) |  |
| High | 48.4 (95% CI, 47.7–49.1) |  |

When it comes to attitudes toward HIV/AIDS, it is noteworthy that almost 46% of the total respondents exhibited a positive attitude (Table 3). Looking at individual components of attitude items, approximately 41% of respondents concurred that children with HIV should not be allowed to attend school alongside other children. Nevertheless, about 54.0% of women expressed their willingness to undergo an HIV test, regardless of concerns about how others might react. Furthermore, more than 60.7% of respondents believed that negative rumours and gossip circulate about people infected with HIV/AIDS and those who live with them. Additionally, around 57.9% of respondents perceived that individuals with HIV/AIDS or those living with an infected person experience a loss of respect and dignity. However, roughly one out of every four respondents admitted to feeling ashamed of residing with someone who has HIV/AIDS.

**Table 3:**
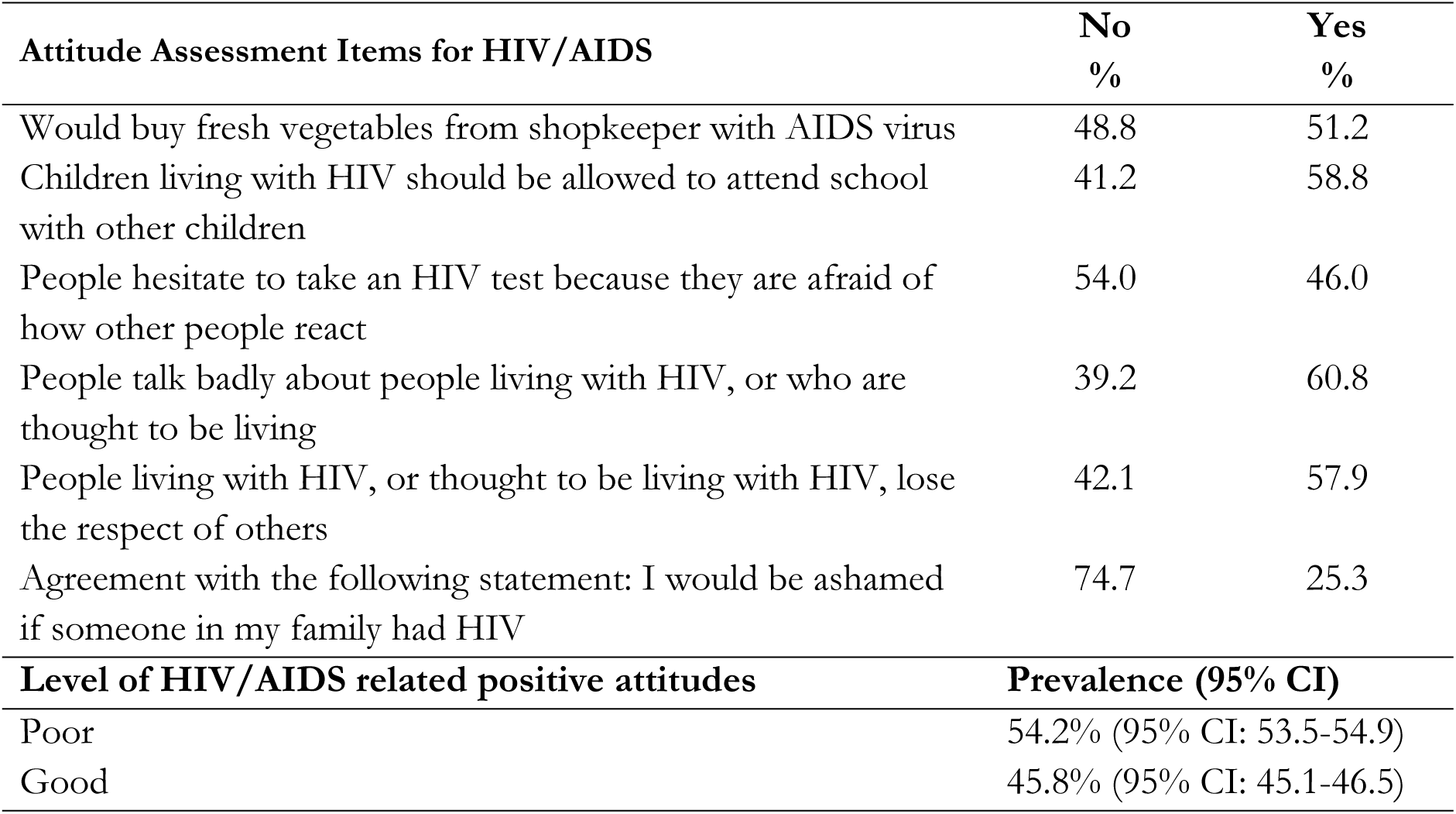
Percentage distribution of items assessing respondents’ attitudes towards HIV/AIDS and composite of positive attitude level (n=39,066)

| Attitude Assessment Items for HIV/AIDS | No<br>% | Yes<br>% |
| --- | --- | --- |
| Would buy fresh vegetables from shopkeeper with AIDS virus | 48.8 | 51.2 |
| Children living with HIV should be allowed to attend school with other children | 41.2 | 58.8 |
| People hesitate to take an HIV test because they are afraid of how other people react | 54.0 | 46.0 |
| People talk badly about people living with HIV, or who are thought to be living | 39.2 | 60.8 |
| People living with HIV, or thought to be living with HIV, lose the respect of others | 42.1 | 57.9 |
| Agreement with the following statement: I would be ashamed if someone in my family had HIV | 74.7 | 25.3 |
| **Level of HIV/AIDS related positive attitudes** | **Prevalence (95% CI)** |  |
| Poor | 54.2% (95% CI: 53.5-54.9) |  |
| Good | 45.8% (95% CI: 45.1-46.5) |  |

### Geographical variations in HIV/AIDS knowledge and attitudes

The geographical distribution of HIV/AIDS knowledge and attitudes is depicted in Figs 1 and 2, respectively. Lower levels of HIV/AIDS knowledge were observed among respondents residing in districts such as Tangail, Munshiganj, Shariatpur, Madaripur, Narail, and Chattogram. Conversely, higher levels of HIV/AIDS knowledge were found among respondents in districts like Sylhet, Lalmonirhat, Rajbari, and Bhola. Regarding attitudes, respondents in districts including Panchagarh, Dinajpur, Kurigram, Lalmonirhat, Jamalpur, Mymensingh, Rajbari, Faridpur, Satkhira, Chandpur, Laksmipur, Bhola, Khagrachari, and Rangamati reported poor attitudes toward HIV/AIDS. Conversely, respondents in districts like Cumilla and Madaripur exhibited good attitudes regarding HIV/AIDS.

**Fig 1.**
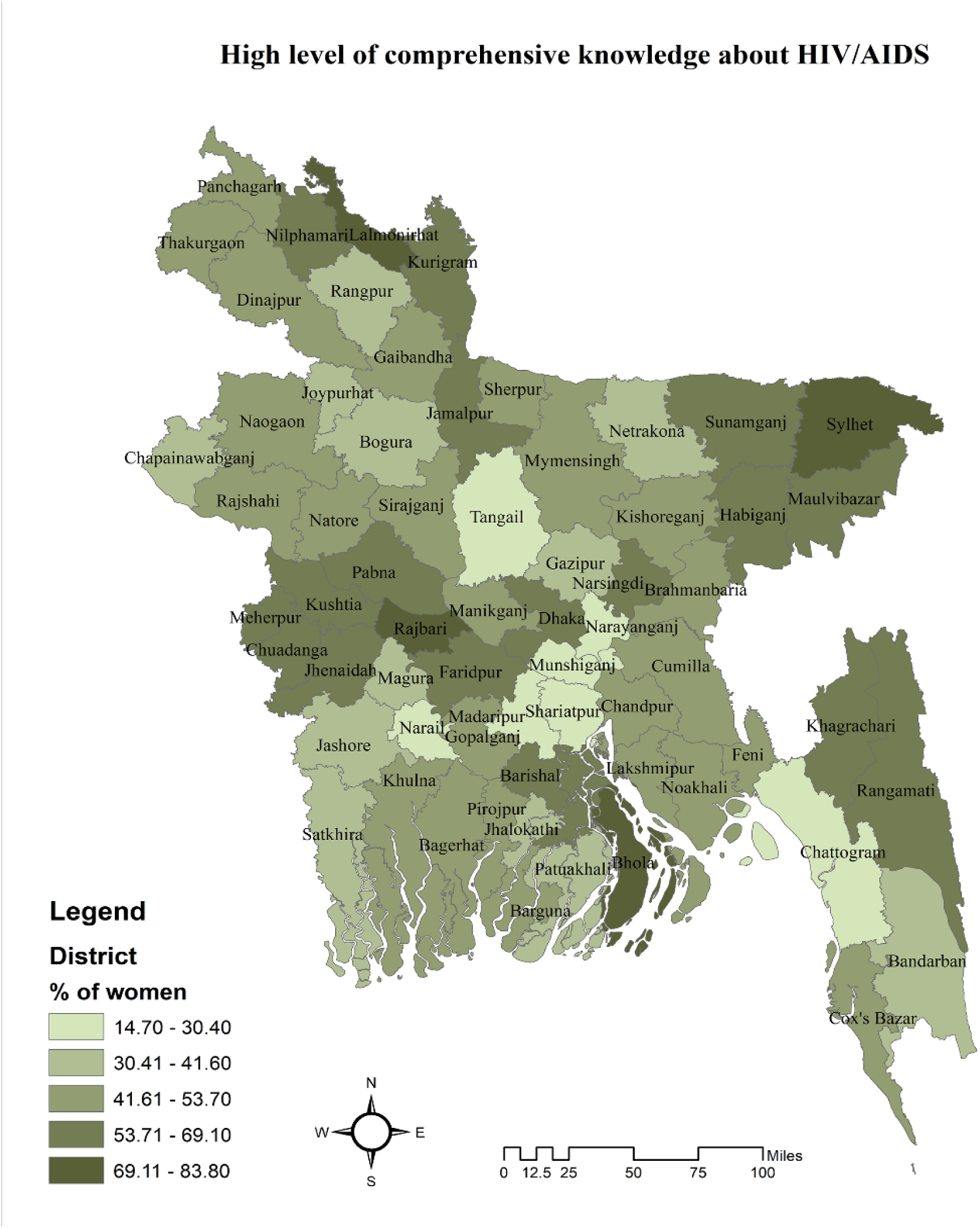
Geographical distribution of high level of comprehensive HIV/AIDS knowledge among reproductive-aged women in Bangladesh (The map was created by the authors using the freely available shape file from https://data.humdata.org)

**Fig 2.**
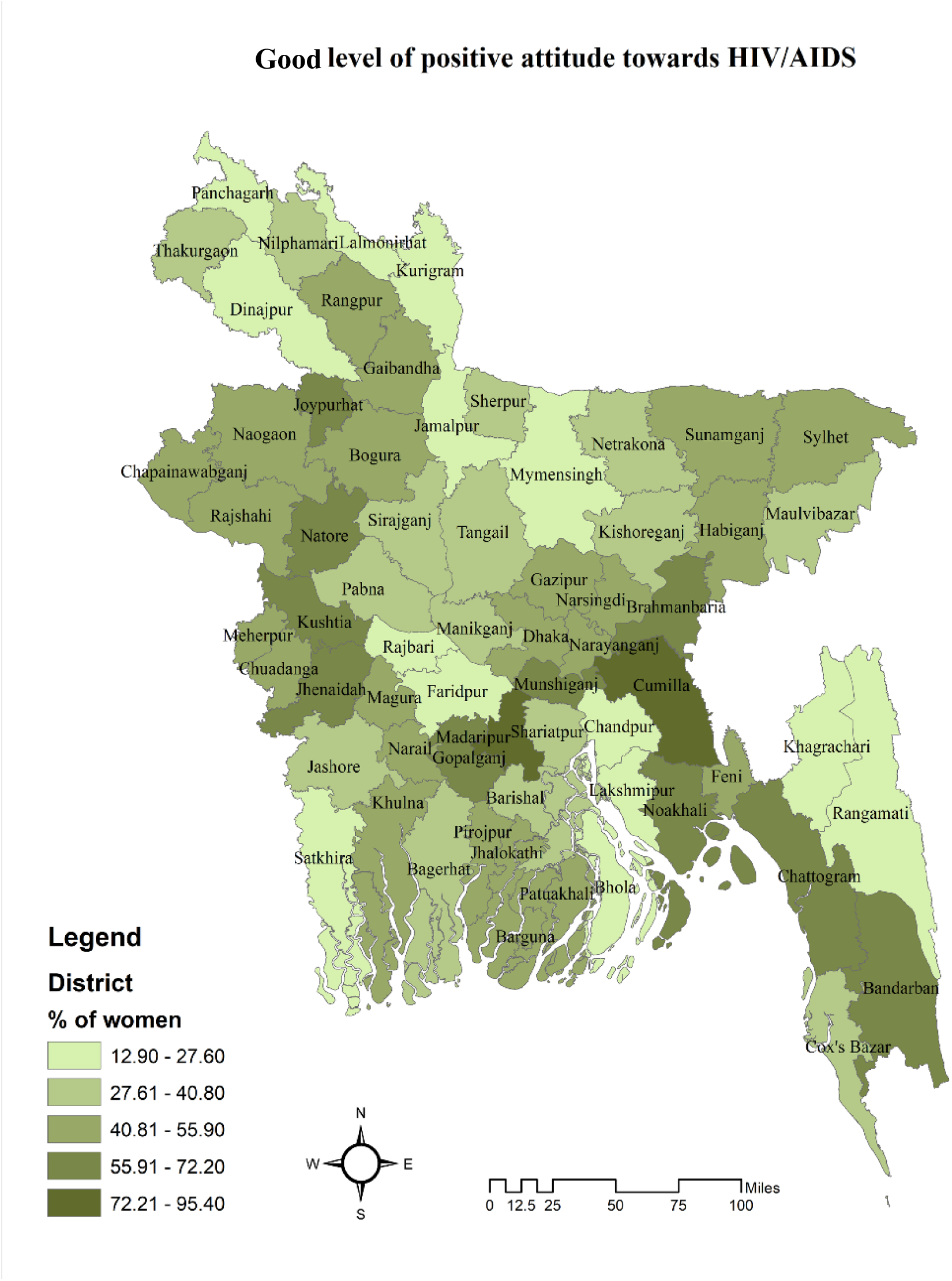
Geographical distribution of good level of positive attitudes regarding HIV/AIDS among reproductive-aged women in Bangladesh (The map was generated by the authors using the freely available shape file from https://data.humdata.org/)

### Multivariate modelling of factors associated with comprehensive HIV/AIDS knowledge and positive attitude towards HIV/AIDS

We employed a multivariate logistic regression model to ascertain the factors linked with a high level of HIV/AIDS knowledge and a positive attitude toward HIV/AIDS, and the findings are presented in Table 4. In comparison to women aged 15-19 years, those in the age groups of 20-24 years (aOR: 1.12, 95% CI: 1.03-1.22, p<0.01), 25-29 years (aOR: 1.16, 95% CI: 1.05-1.27, p<0.01), 30-34 years (aOR: 1.19, 95% CI: 1.08-1.31, p<0.001), 35-39 years (aOR: 1.20, 95% CI: 1.08-1.33, p<0.001), and 40-44 years (aOR: 1.20, 95% CI: 1.07-1.35, p<0.001) were more likely to possess higher HIV/AIDS knowledge. Women who completed lower secondary (aOR: 1.41, 95% CI: 1.25-1.59, p<0.001), secondary (aOR: 2.34, 95% CI: 2.07-2.64, p<0.001), and higher education (aOR: 4.01, 95% CI: 3.45-4.65, p<0.001) had a greater likelihood of having higher HIV/AIDS knowledge compared to women with no formal education. Married women had a 1.09 times (95% CI: 1.0-1.19, p<0.05) greater chance of having higher HIV/AIDS knowledge than unmarried women. Rural women exhibited a 10% lower odds of possessing higher HIV/AIDS knowledge (aOR: 0.90, 95% CI: 0.83-0.96, p<0.01) compared to urban women. Additionally, the likelihood of having higher HIV/AIDS knowledge increased from 1.11 to 1.23 times among respondents residing in households with a better wealth quintile compared to those from poorer households. In comparison to respondents from the Barishal region, those in the Sylhet region had 2.58 times (95% CI: 2.26-2.94, p<0.001) higher odds of reporting higher HIV/AIDS knowledge. Conversely, respondents in the Chattogram (aOR: 0.59, 95% CI: 0.53-0.66, p<0.001), Dhaka (aOR: 0.65, 95% CI: 0.58-0.72, p<0.001), Khulna (aOR: 0.79, 95% CI: 0.71-0.88, p<0.001), Rajshahi (aOR: 0.84, 95% CI: 0.75-0.94, p<0.01), and Mymensingh (aOR: 0.86, 95% CI: 0.75-0.99, p<0.05) regions reported lower likelihoods of higher HIV/AIDS knowledge compared to the respondents in the Barishal region. Women who read newspapers or magazines had 1.27 times (95% CI: 1.16-1.39, p<0.001) greater odds of reporting higher HIV/AIDS knowledge than those who did not. Online media was found to be more effective than mass media. Computer user respondents reported 1.45 times (95% CI: 1.28-1.64, p<0.001) greater odds, internet user respondents reported 1.16 times (95% CI: 1.07-1.25, p<0.001), and mobile user women had 1.13 times (95% CI: 1.06-1.20, p<0.001) greater odds of having a high level of HIV/AIDS knowledge compared to women who were not exposed to these online media-related exposures.

**Table 4:**
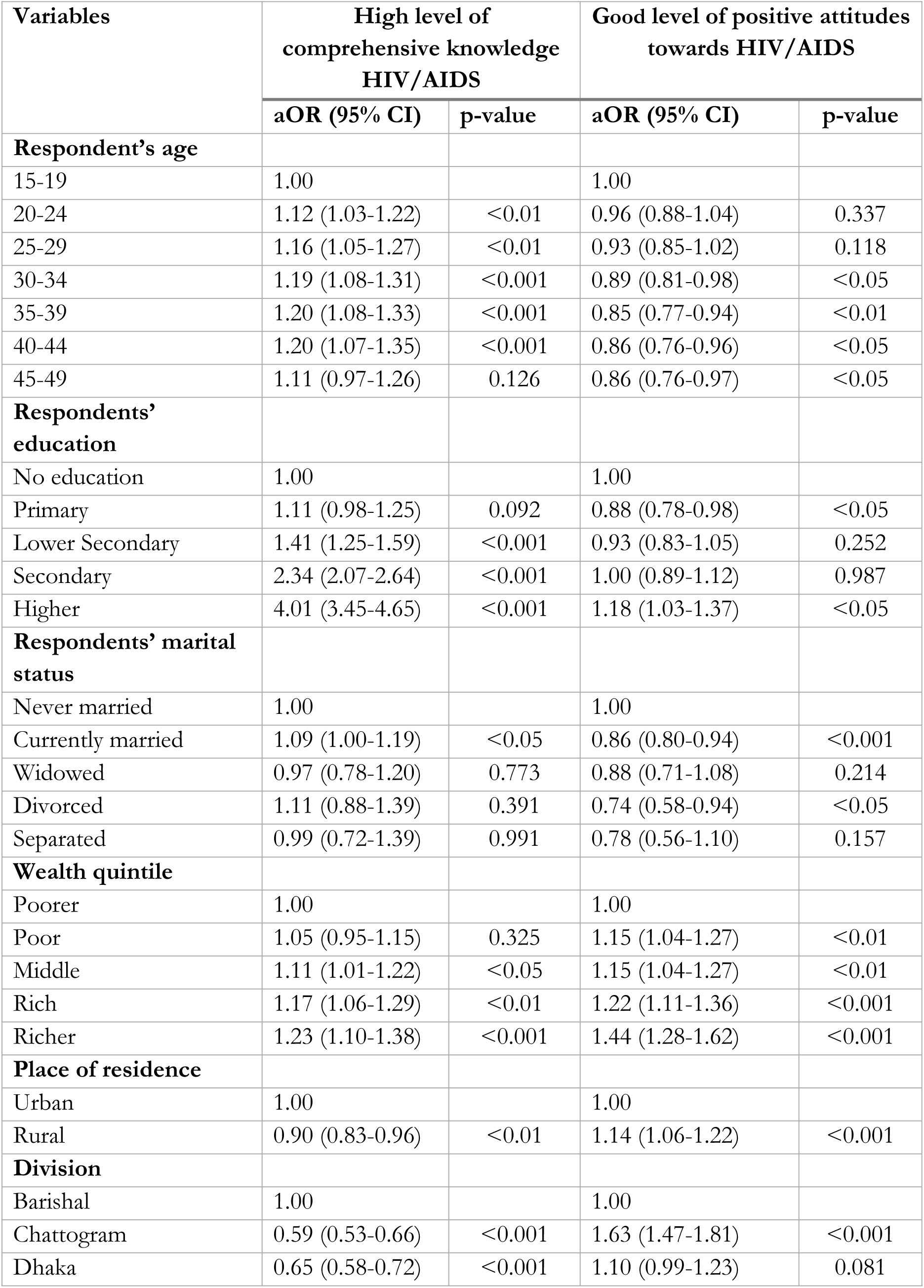
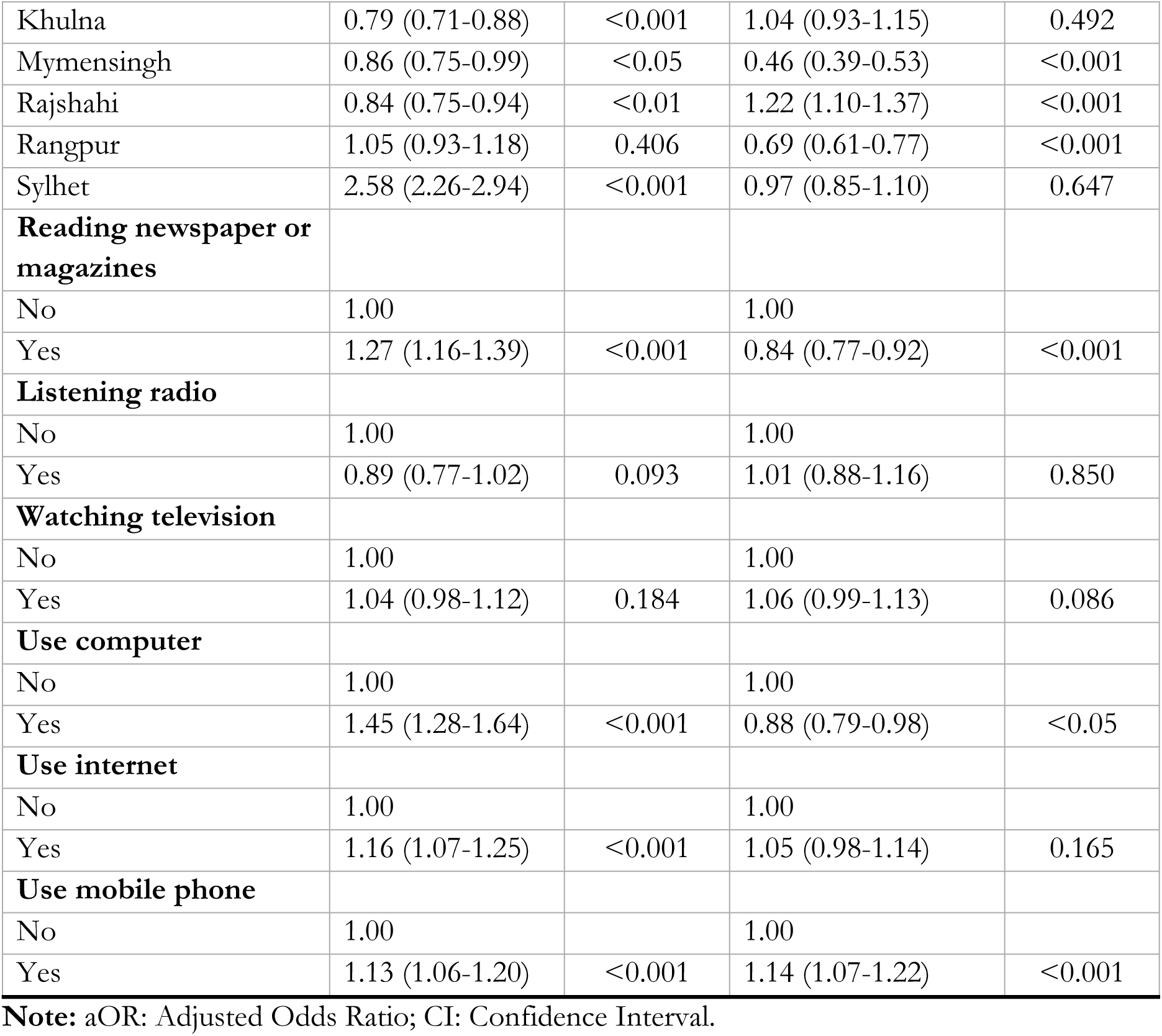
Factors associated with higher level of HIV/AIDS knowledge and positive attitude in Bangladesh: multivariate logistic regression analysis.

We found a declining likelihood of a positive attitude regarding HIV/AIDS among older respondents, with a decrease of 11-15% compared to those aged 15-19. Respondents with higher education had 1.18 times (95% CI: 1.03-1.37, p<0.05) higher odds of a positive attitude toward HIV/AIDS compared to illiterate respondents. Conversely, respondents with primary education (aOR: 0.88, 95% CI: 0.78-0.98, p<0.05) had lower odds of a positive attitude toward HIV/AIDS compared to illiterate respondents. We found a 14% lower likelihood (95% CI: 0.80-0.94, p<0.001) of a positive attitude toward HIV/AIDS among respondents who were currently married compared to never-married respondents. Respondents from poorer to the richest households reported 1.15-1.44 times higher odds of a positive attitude toward HIV/AIDS compared to the poorest households. Rural respondents had 1.14 times more likely to report a positive attitude (95% CI: 1.06-1.22, p<0.001) toward HIV/AIDS compared to urban respondents. Compared to the Barishal region, respondents who lived in Chattogram (aOR: 1.63, 95% CI: 1.47-1.81, p<0.001) and Rajshahi region (aOR: 1.22, 95% CI: 1.10-1.37, p<0.001).

## Discussions

The objectives of this study were to investigate knowledge and attitudes concerning HIV/AIDS, as well as to identify predictors of higher levels of comprehensive knowledge and favorable attitudes toward HIV/AIDS. The findings of this study reveal that over 52% of the total respondents lack good knowledge about HIV/AIDS, and over 54% of the total respondents do not possess a favorable attitude regarding HIV/AIDS. This situation is even more poor in certain districts of Bangladesh. Various socio-demographic and mass-media-related factors were identified as associated with higher levels of knowledge and positive attitudes toward HIV/AIDS. Specifically, higher levels of HIV/AIDS knowledge and favorable attitudes were observed among respondents with higher education levels, residing in households with a comparatively improved wealth quintile, and using mobile phones. Other predictors revealed conflicting associations for knowledge and attitudes regarding HIV/AIDS. For instance, an increasing age, particularly among younger respondents, was significantly associated with higher levels of knowledge about HIV/AIDS but declining favorable attitudes. Additionally, currently married women displayed good knowledge about HIV/AIDS but declining favorable attitudes, while rural women had decreasing probabilities of higher levels of knowledge about HIV/AIDS but higher probabilities of maintaining positive attitudes. These findings are robust, considering the analysis was conducted on a large sample collected from a nationally representative survey, which encompassed a broad range of explanatory variables. The results underscore the need for comprehensive policies and programs aimed at enhancing knowledge and fostering positive attitudes toward HIV/AIDS.

This study reveals a significant lack of sufficient knowledge and poor attitudes regarding HIV/AIDS, with more than half of the women in Bangladesh lacking these attributes, and notable regional variations observed. These findings align with the results of previous studies conducted in Bangladesh [31, 32] and neighboring countries such as India [56, 57] and Pakistan [54]. It is worth noting in general the level of knowledge and attitudes reported in this study can be considered relatively good, given that a majority of the Bangladeshi population has not had direct exposure to individuals affected by HIV/AIDS. Such exposure is an important pathway for increasing awareness and understanding of specific diseases, including HIV/AIDS. However, it is important to recognize that the level of knowledge and attitudes reported in this study pertains solely to reproductive-aged women who have heard about HIV/AIDS. Considering this, and the fact that this group of women in Bangladesh is more likely to have connections with healthcare services due to sexual and reproductive healthcare needs, the findings are concerning. Nevertheless, despite the overall poor conditions, there are a few indicators related to knowledge and attitudes concerning HIV/AIDS that offer hope for improvement. For instance, approximately 75% of the total respondents reported that they would not feel ashamed if a family member had HIV, indicating a potential avenue for enhancing the situation. The reasons for such improvements in specific aspects of knowledge and attitudes are rooted in the strong traditional bonds that Bangladeshi people have with their family members, which tend to outweigh prevalent myths and misconceptions about HIV/AIDS. This situation is also observed in other LMICs [58]. However, the findings of this study regarding knowledge and attitudes about HIV/AIDS also suggest that the national strategic plan in Bangladesh, implemented over the past decades to reduce HIV vulnerabilities, has not been as effective as desired in increasing knowledge and fostering positive attitudes [59].

We also observed variations in knowledge and attitudes regarding HIV/AIDS across several districts in Bangladesh, which are consistent with earlier observations of divisional-level variations in HIV/AIDS knowledge and attitudes in the country [31]. These variations could be attributed to differing levels of mass media presence, variations in educational prevalence at the area level, differences in population distributions, connectivity to healthcare services, and the effectiveness of healthcare service providers in these specific areas [38, 60, 61]. These findings align with previous studies, which indicated that HIV/AIDS-related knowledge and improved attitudes were more prevalent in districts where the government had previously prioritized HIV/AIDS care through hospitals and drop-in centers for HIV/AIDS counseling and treatment. This prioritization was often in response to higher HIV/AIDS prevalence in these districts [7]. These results suggest a potential pathway to enhance HIV/AIDS-related knowledge and attitudes by expanding similar programs implemented in these areas to other regions across Bangladesh.

This study identified several factors contributing to the lower levels of knowledge regarding HIV/AIDS among women in Bangladesh. For instance, rural women reported a reduced likelihood of possessing knowledge about HIV/AIDS and a higher likelihood of maintaining a positive attitude toward the disease. These reported associations can be attributed to multifaceted reasons, with prominent factors including cultural sensitivity and prevailing social norms surrounding sexual issues among rural women. These factors often lead to feelings of shyness and reluctance to openly discuss sexual and reproductive problems [62–64]. Additionally, their lower educational participation, limited engagement outside their rural communities, and limited exposure to mass media, along with reduced use of mobile phones, further exacerbate this issue [30, 65–69]. These findings are consistent with other results of this study that highlight the association of HIV/AIDS knowledge with women’s education and exposure to mass media. It is important to note that these factors, in conjunction with healthcare facility effectiveness and density, can contribute to creating regional variations in HIV/AIDS knowledge [25, 67, 69–72]. These observations align with the findings of this study and are consistent with other research conducted in Bangladesh and other LMICs.

However, the findings of this study, which indicate a higher likelihood of a positive attitude regarding HIV/AIDS among rural women and a lower likelihood of a positive attitude among never-married women, directly contradict the prevailing understanding based on previous research. Earlier studies have reported that urban and unmarried women tend to exhibit more favorable attitudes toward HIV/AIDS [45, 73]. The reasons for these conflicting associations are not entirely clear. However, several factors could contribute to this disparity. One potential explanation could be the stronger family and community-level bonds among rural women in Bangladesh compared to urban women. Additionally, married women in Bangladesh are more likely to access healthcare services, particularly for sexual and reproductive health needs. During these healthcare visits, they may have discussions about HIV/AIDS, which could influence their attitudes toward the disease. Married women may also be more inclined to take precautions to protect themselves and their families, including children, from sensitive sexual issues and diseases, which might lead them to seek knowledge about HIV/AIDS. Moreover, within the cultural norms of Bangladesh, unmarried women often hesitate to discuss sexual health matters with anyone, which significantly influences their attitudes toward HIV/AIDS. Taken together, these factors may contribute to the differences observed between unmarried and married women regarding their attitudes toward HIV/AIDS.

The policy implications of the findings from this study highlight the need to enhance access to awareness-building programs about HIV/AIDS to improve knowledge and attitudes, taking into account the specific needs at the district level. This should be extended to the general population, not limited to women alone, as older generations, in particular, play a crucial role in community-level awareness building. This group, which often holds superstitious beliefs and stigmatizes discussions related to sexuality, is currently not adequately included in existing programs. Textbooks, healthcare facilities, and awareness-building programs can effectively contribute to the generation of knowledge and the shaping of attitudes regarding HIV/AIDS. Although safe sexual and reproductive health-related topics are now part of the secondary education curriculum in Bangladesh, they are not consistently and effectively discussed in the classroom [74–76]. Moreover, women who drop out of primary education miss out on formal channels for acquiring knowledge about HIV/AIDS and developing appropriate attitudes. In addition, door-to-door services aimed at disseminating information about HIV/AIDS are often ineffective. Family planning workers, primarily responsible for these efforts, tend to neglect sharing such knowledge due to their limited numbers, heavy workloads, and lack of monitoring [17, 67, 77]. Addressing these issues can be an effective means of increasing knowledge and attitude toward HIV/AIDS in Bangladesh.

This study has several strengths as well as a few limitations. One of its strengths is the analysis of a nationally representative sample with a large sample size. The data were analyzed using sophisticated statistical methods and included a comprehensive range of explanatory variables. As a result, the findings of this study are robust and can be utilized in the development of national-level programs to increase awareness about HIV/AIDS. However, this study is limited to women who have heard of HIV/AIDS, as the MICS survey only collects knowledge and attitude data from individuals who are aware of it. Additionally, this study excludes men and the hijra population, which limits the generalizability of the findings. Moreover, the data analyzed in this study are cross-sectional in nature, meaning that the reported findings are correlational rather than causal. Furthermore, the data were collected through retrospective questioning, which introduces the risk of recall bias. In addition to the factors considered in this study, it’s important to acknowledge that cultural and traditional norms play a significant role in HIV/AIDS-related knowledge and attitudes. Unfortunately, these aspects were not considered in this study due to their unavailability in the survey. Nevertheless, despite these limitations, the findings generated from this study can be instrumental in shaping national-level policies and programs.

## Conclusions

The study revealed that approximately half of the women in Bangladesh have insufficient knowledge and positive attitudes toward HIV/AIDS, and there are notable regional variations in these aspects. Various socio-demographic factors were identified as associated with higher levels of HIV/AIDS knowledge and favourable attitudes. These findings underscore the urgent need to develop policies and programs aimed at improving HIV/AIDS-related knowledge and attitudes, considering the specific needs at the district level. To achieve this, the utilization of mass media and the integration of HIV/AIDS-related information in textbooks, along with ensuring their effective dissemination and comprehension, can play a pivotal role in enhancing HIV/AIDS-related knowledge and attitudes in Bangladesh.

## Data Availability

The data is openly available to the UNICEF MICS databases in https://mics.unicef.org/surveys.

## Abbreviations

aAOR: adjusted Odd Ratio
ART: Antiretroviral Treatment
BBS: Bangladesh Bureau of Statistics
CI: Confidence Interval
COVID-19: Coronavirus Disease 2019
df: Degrees of Freedom
HIV/AIDS: Human Immunodeficiency Virus/Acquired Immunodeficiency Syndrome
LMICs: Low- and Middle Income Countries
MICS: Multiple Indicator Cluster Survey
PSUs: Primary Sampling Units
PWID: People who inject drugs
SDGs: Sustainable Development Goals
UNICEF: United Nation’s Children Emergency Fund
VIF: Variance Inflation Factor

## Availability of data and materials

The data is openly available to the UNICEF MICS databases in https://mics.unicef.org/surveys.

## Author’s contributors

Conceived and designed the experiments: MAB, RAMY Analyzed the data: MAB; Supervise and Review: RAMY, RMM, MNK; Wrote the paper: MAB, MNK, RAMY, RMM.

## Ethics statement

The data of this study were obtained from Multiple Indicator Cluster Survey (MICS) Archive, supported by United Nation’s Children Fund (UNICEF). The survey protocol was approved by technical committee of the Government of Bangladesh lead by Bangladesh Bureau of Statistics (BBS). Verbal consent was obtained for each respondent participating in the survey. All respondents were informed of the voluntary nature of participation and the confidentiality and anonymity of information and they were informed of their right to refuse answering all or particular questions, as well as to stop the interview at any time.

## Acknowledgements

We thank to the Faculty of Business, Economic and Social Development, Universiti Malaysia Terengganu for conducting the study. Besides we are also acknowledged to the UNICEF for the necessary data, materials and guidelines. Also we are grateful to icddr,b which is grateful to the Government of Bangladesh and Canada for providing core/unrestricted support to icddr,b.

## Supporting information

**S1 table. Comprehensive knowledge items in MICS survey 2019**

**S2 table. Attitude items in MICS survey 2019**

**S1 fig. Sample selection process from the MICS datasets**

**S2 fig. Density function of comprehensive HIV/AIDS knowledge score**

**S3 fig: Density function of attitude towards HIV/AIDS score**

